# Occupational exposure to insulating materials and risk of coronary artery diseases

**DOI:** 10.1101/2022.12.12.22283365

**Authors:** Subhabrata Moitra, Ali Farshchi Tabrizi, Fadi Khadour, Linda Henderson, Lyle Melenka, Paige Lacy

**Author notes:** **Correspondence to:** Paige Lacy, Alberta Respiratory Centre & Division of Pulmonary Medicine, Department of Medicine, 559 Heritage Medical Research Centre, University of Alberta, Edmonton, AB, T6G 2R3, Canada. **Author Contributions:** Conceptualization, L.M.; methodology, F.K., and L.M.; validation, F.K., and L.M.; formal analysis, S.M.; investigation, L.H., F.K., and L.M.; resources, P.L., and L.M.; data curation, A.F.T.; writing—original draft preparation, S.M.; writing—review and editing, all authors; visualization, S.M.; supervision, P.L., L.M.; project administration, L.H., P.L.; funding acquisition, L.M. All authors have read and agreed to the published version of the manuscript.

## Abstract

**Background:** Although previous reports link exposure to insulating materials with an increased risk of mesothelioma and chronic respiratory diseases, studies evaluating their associations with the risk of coronary artery diseases (CAD) are lacking.

**Aims:** We aimed at evaluating associations between exposure to insulating materials and the 10-year risk of CAD among insulators.

**Methods:** In this cross-sectional study, we recruited 643 adults (≥18 years) who were full-time insulators from the Local 110 Heat and Frost Insulators and Allied Workers Union in Edmonton, Alberta. We obtained demographic information, personal and family history, and job-exposure history including experience (years) and types of exposure to insulating materials. Clinical profiling including Framingham risk scores (FRS) was assessed.

**Results:** Of all insulators, 89% were male (mean±SD age: 47±12 years), 27% had a parental history of cardiac diseases, and 22% had a comorbid chronic respiratory disease. 53% reported exposure to asbestos while 61, 82, and 94% reported exposure to ceramic fibers, fiberglass, and mineral fibers, respectively. In single-exposure multivariable regression models adjusted for experience, marital status, and body mass index (BMI), asbestos was found to be associated with higher FRS (β: 1.004; 95%CI: 0.003 to 2.00). The association remained consistent in multi-exposure models and a higher association between asbestos exposure and FRS among insulators with comorbid chronic respiratory disease.

**Conclusions:** Our study demonstrates that apart from cancer and chronic respiratory diseases, asbestos exposure may also have a cardiac effect and thus warranting the need for systematic surveillance to protect workers from the adverse effects of these materials.

**KEY LEARNING POINTS:** *What is already known about this subject:*

- Insulating materials, particularly asbestos and man-made vitreous fibres (MMVFs) are well-known occupational hazards, and both long- and short-term exposure to these materials have been found to associate with increased risk of chronic respiratory diseases and cancers, and mortality due to those diseases.
- Evidence of the cardiovascular effects of asbestos and other MMVFs is scanty and only limited to empirical retrospective mortality studies; however, most of those studies produced mixed results on the roles of asbestos and other MMVFs on cardiovascular health.
- It is important to learn the effects of these insulating materials on organ systems other than the lungs in order to identify the potential health hazards of these materials and upgrade surveillance and current safety measures to protect the workers.

*What this study adds:*

- We studied over 800 workers who were exposed to a variety of insulating materials including asbestos and MMVFs and our results indicate an association between asbestos exposure and increased Framingham risk score (10-year estimated risk of cardiovascular diseases).
- Further long-term prospective studies are needed to assess the short and long-term effects of these exposures on disease development, particularly among non-smokers. Furthermore, the quantitation of workplace exposure is also important to understand the dose-response nature of these exposures.

*What impact this may have on practice or policy:*

- Our findings provide new evidence of asbestos-associated increased risk of cardiovascular diseases, which underscores the need for a more rigorous systematic monitoring of the health of workers.
- Our findings also warrant a more comprehensive knowledge of the physicians, particularly occupational physicians regarding the possible occupational risk factors for chronic diseases, including cardiovascular diseases.
- Our findings may provide a basis for further research and amendment of policies related to the workers’ health, protection, compensation, and benefits.

**TEASER TEXT:** Previous studies have demonstrated that exposure to insulating materials including asbestos is associated with an increased risk of mesothelioma and other chronic respiratory diseases. However, these materials have not been systematically investigated as possible risk factors for coronary artery diseases (CAD). Our study is the first to demonstrate an association between exposure to insulating materials, particularly asbestos, and an increased 10-year predicted risk of CAD. These findings warrant further control of exposure to these materials.

## INTRODUCTION

Several new man-made materials, often known as man-made vitreous fibres (MMVFs), aerogels, carbon fibers, mineral fibers, and refractory ceramic fibers have been introduced as alternative insulating materials after many countries imposed a ban on the use of asbestos. Most of these fibres also possess significant health hazards, particularly affecting the respiratory system leading to numerous adverse conditions such as chronic chest infection, carcinoma, adverse pleural conditions, and to some extent, obstructive and interstitial changes in the lungs [1-10]. However, despite the ban on asbestos use, workers, particularly construction workers and insulators continue to be exposed to these materials, for example, during demolition, removal, or renovation of old constructions, insulation, etc.

Although seminal information about the toxicity and possible adverse respiratory health effects of these materials are available from historic and contemporary clinical and public health studies, a majority of those largely focused on respiratory and cancer-related outcomes, and little is known about their involvement in other target organs, e.g., cardiovascular health. While some previous studies indicated possible cardiovascular health effects of occupational asbestos exposure, most of them have been primarily investigated post-mortem, i.e., the involvement of asbestos in cardiovascular disease-related mortality could only be established retrospectively [11, 12]. Nevertheless, several reports showed no significant cardiovascular effects regarding asbestos exposure [13]. Lastly, many of those studies also reported the co-occurrence of malignancies and other chronic diseases because of asbestos exposure, therefore, whether such adverse cardiovascular health conditions were caused by asbestos, or the consequences of other health conditions was not clear. Similarly, literature on the possible cardiovascular health effects of MMVFs is scarce and produce mixed evidence. However, whether exposure to such materials is specifically associated with the risk of coronary artery disease (CAD) has not been systematically investigated.

In this study, we aimed to investigate whether occupational exposure to insulating materials (asbestos, aerogels, calcium silicate, carbon fibers, fiberglass, mineral fibers, and refractory ceramic fibers) is associated with an increased risk of CAD in insulators.

## METHODS

In this cross-sectional study, we investigated 843 unionized insulators of the Local 110 Heat and Frost Insulators and Allied Workers Union in Edmonton, Alberta. All participants were screened at Synergy Respiratory Care Clinic, Sherwood Park, Alberta. Details of the study design, inclusion and exclusion criteria, and methodologies have been reported elsewhere [1, 2]. In brief, the participants were administered a questionnaire containing items of demographic profile, personal and family history (smoking history and pack-years, alcohol consumption, parental history of any cardiac diseases, and frequency of weekly physical activity), and detailed job-exposure history including experience (years), types of exposure to insulating materials, and use of personal protective equipment (PPE) at work for each of the materials (such as aerogels, asbestos, calcium silicate, carbon fibers, fiberglass, mineral fibers, and refractory ceramic fibers [RCFs]).

Detailed clinical profiling of the participants was performed at the clinic that included an assessment of any current respiratory and cardiac conditions such as hypertension and chronic respiratory diseases and any previous incidents of cardiorespiratory or metabolic conditions such as chest pain/angina, heart attack, heart failure, heart failure, cardiac catheterization, coronary bypass surgery, angioplasty, atrial fibrillation, lung cancer, diabetes mellitus, or any other acute conditions. Current medication status was obtained from questionnaires and was verified from the participants’ health records. A venous blood sample was collected, and the plasma lipid profiles (cholesterol, triglyceride, high and low-density lipoproteins) were analysed. Framingham risk score (the 10-year risk of manifesting clinical cardiovascular diseases such as coronary artery diseases [CAD], stroke, peripheral vascular diseases [PVD], chronic heart failure [CHF], and cardiac death) was calculated according to previously established formulae [14, 15].

Data were presented as mean (standard deviation [SD]), median (interquartile range [IQR]), or frequency (%) for continuous, ordinal, and categorical variables, respectively. We first tested the bivariate relationships between each of the exposures (yes/no) and Framingham risk score (FRS) using Student’s t-tests. We then constructed univariable (unadjusted) and multivariable (adjusted) regression models for each exposure and FRS using linear regression models. Ethnicity, education, marital status, body mass index [BMI], years of exposure to insulation work, and the use of PPE were tested as potential confounders. As age, sex, and smoking status were adjusted while calculating the FRS, they were not further considered confounders. Models were constructed in step-forward and step-backward algorithms and only marital status, body mass index (BMI), and years of exposure to insulation works were retained in the final models based on the Akaike information criterion (AIC) for model selection [16].

We also performed several secondary analyses. Firstly, we created a multi-exposure linear model (taking all the exposures in a single model) and tested the associations with FRS as the exposure variables did not demonstrate collinearity among them (variance inflation factor, VIF <2). Secondly, we stratified the single-exposure models by sex and tested the pair-wise differences of the coefficients using the Chow test [17]. Lastly, we tested potential effect modification by alcoholic drink per month (<1 vs. ≥1 per month), any physical activity other than regular work (no/yes), parental history of cardiac disease (no/yes), and any chronic respiratory disease (no/yes). All analyses were conducted using a complete case approach in Stata V.17 (StataCorp, College Station, TX, USA), and a p-value < 0.05 was considered statistically significant.

The present study has been conducted as per the Declaration of Helsinki and is compliant with the Strengthening the Reporting of Observational Studies in Epidemiology (STROBE) guidelines [18], and approved by the Health Research Ethics Board of Alberta (HREBA.CTC-17-0067) and Health Research Ethics Board (Pro00079792), University of Alberta. All participants provided signed informed consent forms before taking part in the study.

## RESULTS

Of those 843 insulators who were screened, we excluded those with a current or previous clinical diagnosis of one or more conditions such as chronic chest pain/angina (n=119), heart attack (n=20), heart failure (n=3), cardiac catheterization (n=22), coronary bypass (n=8), angioplasty (n=21), atrial fibrillation (n=27), mesothelioma (n=1), or other chronic or acute cardiopulmonary conditions (n=118). We finally recruited 643 insulators for this analysis of which 571 (89%) were male with a mean age (SD) of 47 (12) years. 78% were Caucasians, 67% were ever smokers, 27% reported a family history of cardiac diseases, and 22% had a comorbid chronic respiratory disease (**Table 1**). Exposure to different insulating materials ranged between 39 (aerogels) - 94% (refractory ceramic fiber) and 53% of the workers reported having exposure to asbestos at workplaces. The mean (SD) FRS was 7.3 (6.6).

**Table 1:** Demographic profile, exposure history and clinical profile of the insulators

|  | **N = 643** |
| --- | --- |
| Age (years), mean (SD) | 47 (12) |
| Sex (male), n (%) | 571 (89) |
| BMI (Kg/m<sup>2</sup>), mean (SD) | 30 (15) |
| Ethnicity (Caucasian), n (%) | 501 (78) |
| Education, n (%) |  |
| Grade school | 17 (3) |
| High school | 77 (12) |
| Trade school | 302 (47) |
| College/university | 247 (38) |
| Marital status (married), n (%) | 407 (63) |
| Smoking status, n (%) |  |
| Ever smoker | 429 (67) |
| Overall pack-years, median (IQR) | 5 (0 – 17) |
| Any chronic respiratory diseases, n (%) | 141 (22) |
| Alcohol consumption, n (%) |  |
| < 1 drink per month | 77 (12) |
| Physical activity, n (%) |  |
| Never | 297 (46) |
| 1-3 times/week | 209 (33) |
| 4-6 times/week | 65 (10) |
| Daily | 72 (11) |
| Parental history of cardiac disease, n (%) | 169 (27) |
| ***Exposure variables*** |  |
| Years of exposure in insulation work, median (IQR) | 13 (4 – 28) |
| Exposure to insulating materials, n (%) |  |
| Aerogels | 253 (39) |
| Asbestos | 343 (53) |
| Calcium silicate | 552 (86) |
| Carbon fibers | 370 (58) |
| Ceramic fibers | 393 (61) |
| Fiberglass | 525 (82) |
| Mineral fibers | 603 (94) |
| PPE used for, n (%) |  |
| Aerogels | 227 (35) |
| Asbestos | 181 (28) |
| Fiberglass | 282 (44) |
| Mineral fibers | 447 (70) |
| ***Cardiometabolic variables*** |  |
| Systolic blood pressure (mmHg), median (IQR) | 124 (115 – 132) |
| Clinically diagnosed hypertension, n (%) | 117 (18) |
| Medications for hypertension, n (%) | 91 (14) |
| Lipid profile (mg/dL), mean (SD) |  |
| Cholesterol | 195.2 (41.5) |
| Triglyceride | 140.2 (90.2) |
| HDL | 50.8 (14.6) |
| LDL | 118.1 (36.0) |
| Framingham risk score, mean (SD) | 7.3 (6.6) |

Data presented as mean (standard deviation [SD]), median (interquartile range [IQR]), or frequencies (%) as described in the text.
Abbreviations: BMI: body mass index; HDL: high density lipoprotein; LDL: low density lipoprotein; PPE: personal protective equipment

In single-exposure univariable (unadjusted) models, asbestos, ceramic fibres, and fibreglass (β range: 1.28 to 4.66; all p-values < 0.05) were associated with higher FRS (**Figure 1A & Supplementary Table 1**); however, in the adjusted (multivariable) models, only asbestos retained significance association with higher FRS (β: 1.004; 95%CI: 0.003 to 2.00). In the multi-exposure model, only asbestos remained associated with higher FRS (β: 1.08; 95%CI: 0.05 to 2.10) (**Figure 1B & Supplementary Table 1**).

**Figure 1:**
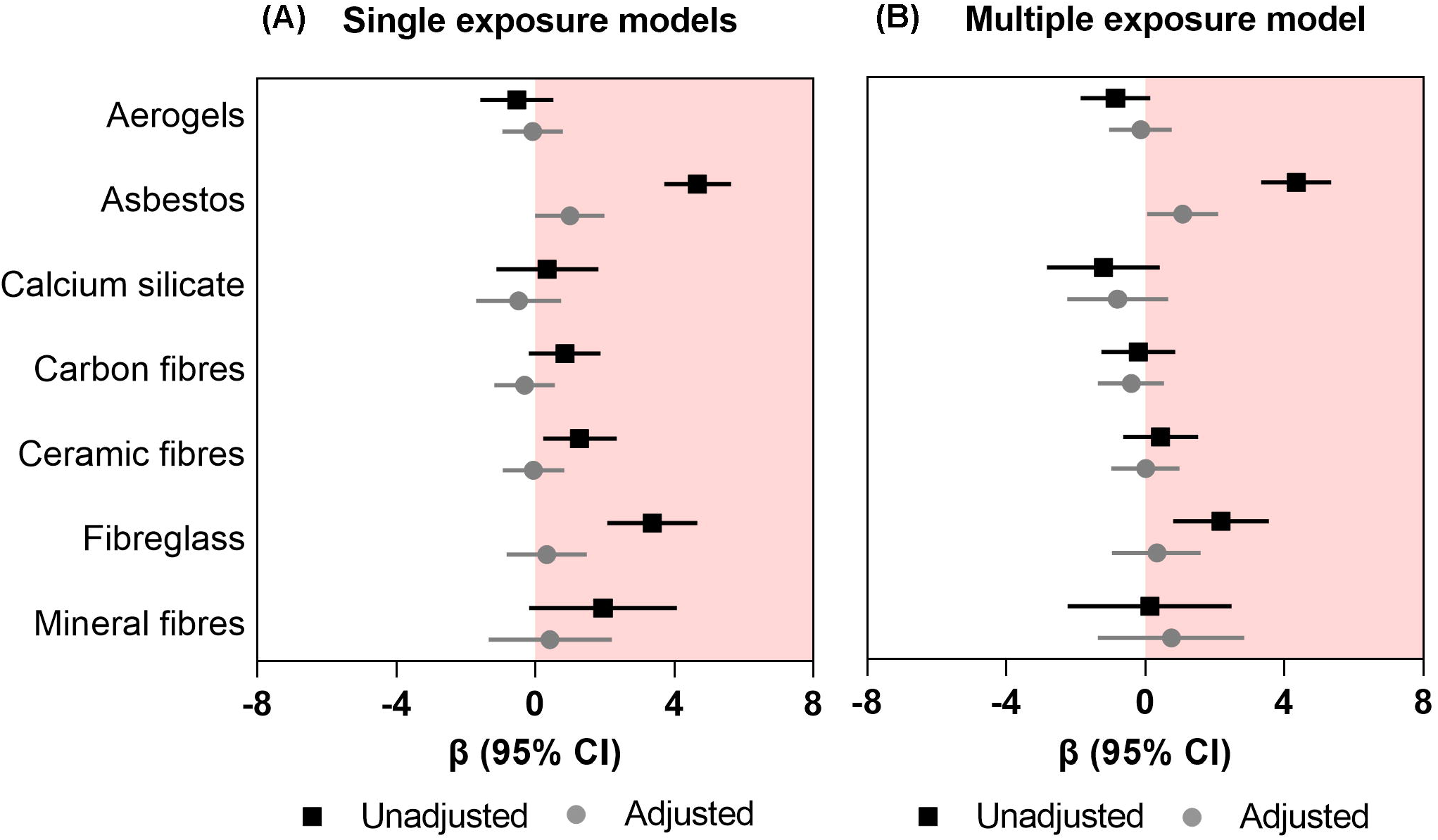
Association between occupational exposure to mineral fibers and Framingham risk score (A) Single-exposure models (B) Multi-exposure models. Data presented as regression coefficient (marker) and 95% confidence interval (error bars) adjusted for marital status, BMI, and years of exposure in insulation works.

In the secondary analyses, we observed a higher association between asbestos exposure and FRS in male insulators (β: 1.14; 95%CI: 0.06 to 2.23) than their female counterpart (β: -0.18; 95%CI: - 1.31 to 0.97); however, the difference of the coefficients was not significant (chi-squared p-value for Chow test = 0.13) (**Figure 2A & Supplementary Table 2**). Furthermore, we found that the association between asbestos exposure and FRS was higher among the insulators who had a comorbid chronic respiratory disease (β: 3.05; 95%CI: 0.61 to 5.50) than those without any comorbid chronic respiratory diseases (β: 0.36; 95%CI: -0.72 to 1.43; p-value for interaction: 0.007) (**Figure 2B & Supplementary Table 3**). We did not observe any clinically important influence of other effect modifiers (alcoholic drink per month, any physical activity other than regular work, or parental history of cardiac disease) on the associations between insulating materials and FRS (**Supplementary Tables 4-6**).

**Figure 2:**
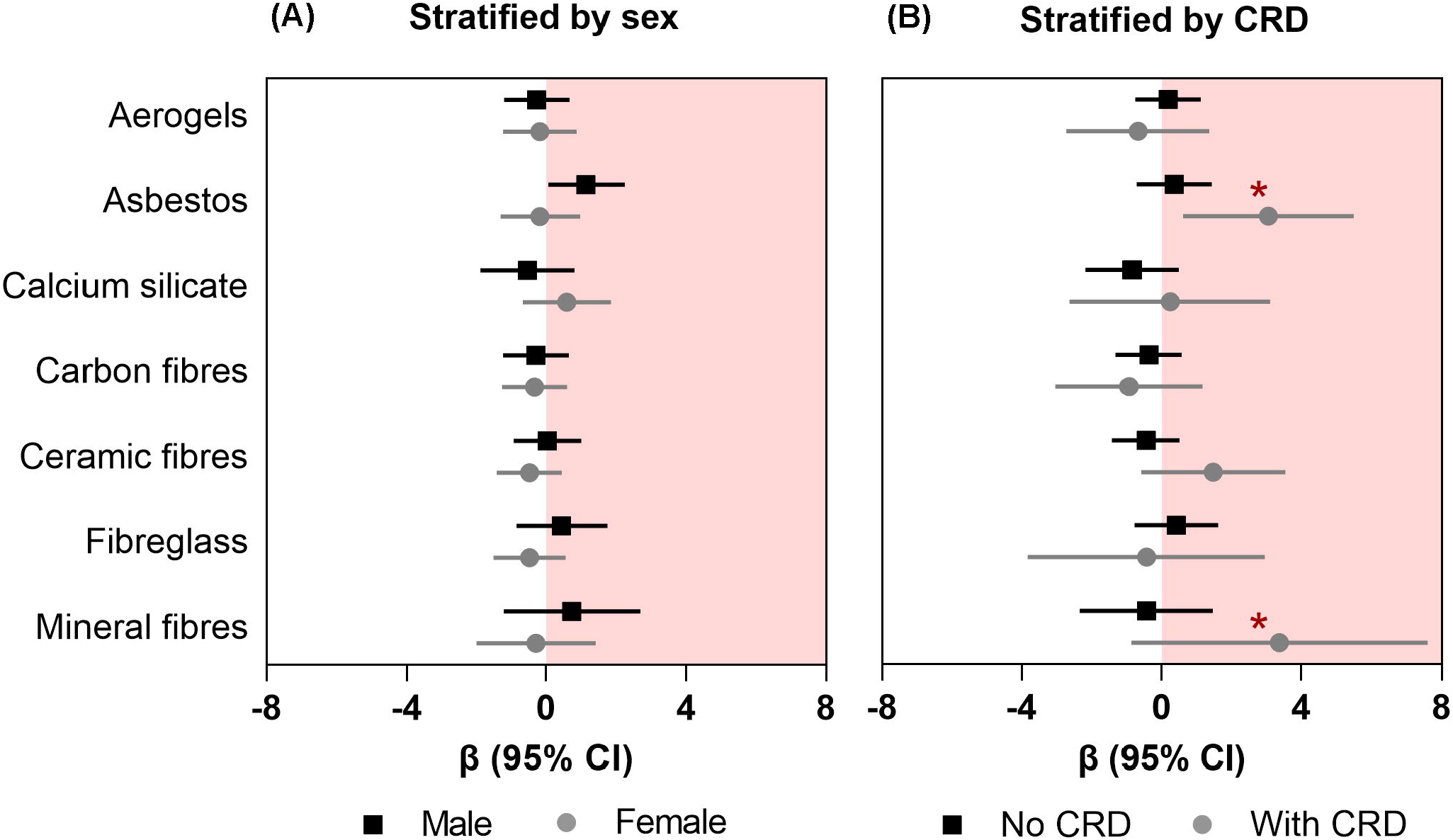
Association between occupational exposure to mineral fibers and Framingham risk score (A) stratified by sex and (B) effect modification by any chronic respiratory disease (CRD) Data presented as regression coefficient (marker) and 95% confidence interval (error bars) adjusted for marital status, BMI, and years of exposure in insulation works. * indicates the p-values for interaction that are significant at p<0.05.

## DISCUSSION

To our knowledge, this is the first prospective study assessing the relationship between several types of insulating materials and the 10-year predictive risk score for CAD in a group of workers. We observed that of all insulating materials, only asbestos exposure was associated with a higher Framingham risk score. This association was stronger among males and those with comorbid chronic respiratory disease. We did not find any associations between other insulating materials and Framingham risk score or any influence of possible behavioural or genetic factors such as alcohol consumption, physical activity, or family history of cardiac diseases.

Our findings of asbestos-associated increased projected risk of cardiovascular diseases extend the observations of a recent meta-analysis where Rong et al. (2015) demonstrated the pooled standardized mortality ratio (SMR) estimate for cardiovascular-related diseases of 1.11 (95% CI: 1.01-1.22), indicating a significant association between asbestos exposure and an increased risk of cardiovascular-related diseases in exposed workers [19]. Nevertheless, despite numerous post-mortem studies on asbestos exposure and cardiovascular disease-related mortality, the mechanisms of asbestos exposure-associated cardiovascular diseases have not been fully understood. One possible explanation could be the physiological alteration of the heart and the vasculature due to asbestos exposure as a previous study showed pericardial thickening in asbestos-exposed workers [20]. Another possible explanation for the increased risk of cardiovascular diseases could be the inflammatory effects of asbestos on the cardiopulmonary circuit, leading to a sustained accumulation of inflammatory mediators, and an upsurge in oxidative stress, all of which possibly help in the development of atherosclerotic plaques [21]. While some animal studies support this hypothesis of free radical production, inflammation in coronary arteries and vascular and thrombogenic effects of asbestos exposure [22-24], human data on the mechanisms are unavailable and difficult to obtain.

Our findings of the roles of other insulating MMVFs on FRS are similar to the meta-findings of previous studies [25-31], which did not indicate any significant associations between MMVFs and cardiovascular diseases. Although a relatively recent study reported a higher increased risk for ischemic heart disease among Swedish construction workers, the study used a job-exposure matrix that considered a wide array of exposures such as dust, gas, fumes, and other particulate matters [32]. Therefore, it was not clear whether the increased risk of cardiovascular diseases was specifically attributed to exposure to MMVFs.

One of the major strengths of this study is its novel approach to assessing a chronic and probable detrimental cardiological effect of the insulating materials that have not been previously investigated. Secondly, we considered a wide range of insulating materials and tested their individual and cumulative associations with the FRS. Lastly, we performed several secondary analyses to test the plausible influence of other potential lifestyle and clinical factors on the association between insulating materials and FRS, which highlighted the significant influence of comorbid chronic respiratory diseases. However, the study has some limitations as well. As the current analysis is cross-sectional in design, we could not determine any causal association. Secondly, we only had information on whether the insulators were exposed to insulating materials or not, and we could not measure the level of exposure to each insulating material or determine the cumulative exposure index. Similarly, information about PPE (duration of use, brand, or specifications) was not available. However, adjusting the models for PPE did not significantly alter the magnitude of the estimates, which means that the PPE did not provide substantial protection to the workers from exposure. Moreover, we could not perform any detailed biochemical studies, such as oxidative stress assessment, which could have been useful to explain the cardiological impacts of these insulating materials. Lastly, we did not have detailed information about other potential workplace exposures such as physical and chemical agents, heat or cold exposure, and other behavioural and socioenvironmental triggers, which could also impart adverse health effects. Thus, these factors also need to be considered in future studies.

Our findings highlight an important health concern of asbestos exposure and recommend more holistic surveillance of the workplaces, amend adequate protective measures, and administer more frequent and rigorous monitoring of health to minimize the risk of work-related health hazards. Asbestos-related diseases are often diagnosed years after such exposures due to their long latency. Therefore, it is important for physicians to consider possible work-related exposures to hazardous materials which might influence the development of such diseases.

We may conclude that occupational exposure to insulation materials, particularly asbestos is associated with a higher risk of CAD in insulators and the risk is higher among those who have a comorbid chronic respiratory disease such as asthma or COD. While the adaptation of less hazardous insulating materials and the use of proper PPE are recommended, a regular health monitoring and workplace surveillance program must be implemented to diagnose.

## Data Availability

All data produced in the present study are available upon reasonable request to the authors.

## Funding & Acknowledgements

This research was Supported by grants from the Wellness of Workers (WoW) Program, Local 110 Heat & Frost Insulators & Allied Workers, and Synergy Respiratory Care Limited. However, the funder was not involved in the design of the study and did not influence the dissemination of the study outcomes. The authors thank all the volunteer insulators contributing to the WoW survey program as well as members of Synergy Respiratory and Cardiac Care: Dina Fathy, Bill Stowe, Carrie McPhee, Jennifer Spring, Melanie De Pruis, Lindsay Simmonds, Celeste Hernandez, Kylie Haydey, Blake MacDonald, Linda Ferguson, Michael Krause, Heather Ryan, Michele McKim, Alana Goodall, Shirley Carr, Kevin Lecht, William Spring, and Melanie Holowach. We would also like to thank Meghan Dueck and Lei Pei for their support in data management.

