## Supplementary Tables for "Occupational exposure to insulating materials and risk of coronary artery diseases": Supplementary tables.docx

**Supplementary Table 1:** Association between exposure to insulating materials and Framingham risk score

**Supplementary Table 2:** Association between exposure to insulating materials and Framingham risk score stratified by sex

**Supplementary Table 3:** Effect modification by any chronic respiratory disease (CRD) (no vs. yes) on the association between exposure to insulating materials and Framingham risk score.

**Supplementary Table 4:** Effect modification by alcoholic drink per month (<1 vs. ≥1 per month) on the association between exposure to insulating materials and Framingham risk score.

**Supplementary Table 5:** Effect modification by physical activity (no vs. yes) on the association between exposure to insulating materials and Framingham risk score.

**Supplementary Table 6:** Effect modification by parental history of cardiac disease (no vs. yes) on the association between exposure to insulating materials and Framingham risk score.

**Supplementary Table 1:** Association between exposure to insulating materials and Framingham risk score

| **Exposure** | **Single exposure model [β (95% C)]** | | **Multiple exposure model [β (95% C)]** | |
| --- | --- | --- | --- | --- |
|  | **Unadjusted** | **Adjusted** | **Unadjusted** | **Adjusted** |
| Aerogels | -0.53 (-1.58 to 0.53) | -0.07 (-0.94 to 0.80) | -0.85 (-1.85 to 0.14) | -0.13 (-1.02 to 0.76) |
| Asbestos | **4.66 (3.70 to 5.63)** | **1.004 (0.003 to 2.00)** | **4.35 (3.34 to 5.36)** | **1.08 (0.05 to 2.10)** |
| Calcium silicate | 0.35 (-1.12 to 1.83) | -0.48 (-1.70 to 0.75) | -1.19 (-2.82 to 0.42) | -0.79 (-2.23 to 0.66) |
| Carbon fibres | 0.86 (-0.18 to 1.89) | -0.30 (-1.18 to 0.57) | -0.20 (-1.25 to 0.86) | -0.41 (-1.35 to 0.54) |
| Ceramic fibres | **1.28 (0.23 to 2.33)** | -0.05 (-0.93 to 0.84) | 0.44 (-0.64 to 1.52) | 0.01 (-0.96 to 0.98) |
| Fibreglass | **3.36 (2.06 to 4.66)** | 0.33 (-0.82 to 1.49) | **2.18 (0.80 to 3.56)** | 0.33 (-0.94 to 1.59) |
| Mineral fibres | 1.95 (-0.17 to 4.07) | 0.43 (-1.34 to 2.19) | 0.13 (-2.22 to 2.49) | 0.75 (-1.35 to 2.85) |

Data presented as regression coefficient (β) and 95% confidence interval (CI) unless otherwise stated. In adjusted models, marital status, body mass index (BMI), and years of exposure to insulation works were considered as confounders.

**Supplementary Table 2:** Association between exposure to insulating materials and Framingham risk score stratified by sex.

| **Exposure** | **Framingham risk score [β (95% C)]** | | ***P*^†^** |
| --- | --- | --- | --- |
|  | **Male (n = 571)** | **Female (n = 72)** |  |
| Aerogels | -0.27 (-1.21 to 0.67) | -0.18 (-1.24 to 0.87) | 0.79 |
| Asbestos | **1.14 (0.06 to 2.23)** | -0.18 (-1.31 to 0.97) | 0.13 |
| Calcium silicate | -0.54 (-1.89 to 0.81) | 0.59 (-0.67 to 1.84) | 0.22 |
| Carbon fibres | -0.29 (-1.24 to 0.65) | -0.33 (-1.27 to 0.60) | 0.60 |
| Ceramic fibres | 0.03 (-0.93 to 1.00) | -0.48 (-1.42 to 0.45) | 0.50 |
| Fibreglass | 0.44 (-0.85 to 1.73) | -0.48 (-1.51 to 0.56) | 0.43 |
| Mineral fibres | 0.73 (-1.22 to 2.68) | -0.29 (-2.00 to 1.42) | 0.45 |

Data presented as regression coefficient (β) and 95% confidence interval (CI) unless otherwise stated. Models were adjusted for marital status, body mass index (BMI), and years of exposure to insulation works were considered as confounders. *P*^†^: chi-squared p-values for Chow test

**Supplementary Table 3:** Effect modification by any chronic respiratory disease (CRD) (no vs. yes) on the association between exposure to insulating materials and Framingham risk score.

| **Exposure** | **Framingham risk score [β (95% C)]** | | ***P^#^*** |
| --- | --- | --- | --- |
|  | **No CRD (n = 502)** | **With CRD (n = 141)** |  |
| Aerogels | 0.18 (-0.76 to 1.12) | -0.68 (-2.72 to 1.36) | 0.40 |
| Asbestos | 0.36 (-0.72 to 1.43) | **3.05 (0.61 to 5.50)** | **0.007** |
| Calcium silicate | -0.84 (-2.17 to 0.49) | 0.24 (-2.63 to 3.10) | 0.24 |
| Carbon fibres | -0.37 (-1.31 to 0.57) | -0.93 (-3.03 to 1.17) | 0.82 |
| Ceramic fibres | -0.45 (-1.41 to 0.51) | 1.47 (-0.59 to 3.54) | 0.07 |
| Fibreglass | 0.42 (-0.78 to 1.61) | -0.44 (-3.82 to 2.95) | 0.87 |
| Mineral fibres | -0.44 (-2.33 to 1.46) | 3.37 (-0.87 to 7.60) | **0.03** |

Data presented as regression coefficient (β) and 95% confidence interval (CI) unless otherwise stated. Models were adjusted for marital status, body mass index (BMI), and years of exposure to insulation works were considered as confounders. *P*^#^: p-values for interaction

**Supplementary Table 4:** Effect modification by alcoholic drink per month (<1 vs. ≥1 per month) on the association between exposure to insulating materials and Framingham risk score.

| **Exposure** | **Framingham risk score [β (95% C)]** | | ***P^#^*** |
| --- | --- | --- | --- |
|  | **< 1 drink per month (n = 77)** | **≥ 1 drink per month (n = 565)** |  |
| Aerogels | **-3.64 (-6.65 to -0.64)** | 0.33 (-0.58 to 1.23) | **0.02** |
| Asbestos | 2.34 (-1.52 to 6.21) | 0.75 (-0.28 to 1.78) | 0.34 |
| Calcium silicate | -0.38 (-4.25 to 3.48) | -0.47 (-1.77 to 0.82) | 0.94 |
| Carbon fibres | -0.08 (-3.01 to 2.85) | -0.28 (-1.19 to 0.64) | 0.93 |
| Ceramic fibres | 1.44 (-1.59 to 4.48) | -0.21 (-1.14 to 0.71) | 0.30 |
| Fibreglass | 3.20 (-1.65 to 8.04) | 0.001 (-1.17 to 1.18) | 0.11 |
| Mineral fibres | 1.01 (-4.58 to 6.60) | 0.30 (-1.56 to 2.17) | 0.70 |

Data presented as regression coefficient (β) and 95% confidence interval (CI) unless otherwise stated. Models were adjusted for marital status, body mass index (BMI), and years of exposure to insulation works were considered as confounders. *P*^#^: p values for interaction.

**Supplementary Table 5:** Effect modification by physical activity (no vs. yes) on the association between exposure to insulating materials and Framingham risk score.

| **Exposure** | **Framingham risk score [β (95% C)]** | | ***P^#^*** |
| --- | --- | --- | --- |
|  | **No physical activity (n = 297)** | **Physical activity (n = 346)** |  |
| Aerogels | 0.05 (-1.27 to 1.36) | -0.20 (-1.35 to 0.95) | 0.86 |
| Asbestos | 0.72 (-0.82 to 2.26) | 0.99 (-0.31 to 2.29) | 0.67 |
| Calcium silicate | -0.39 (-2.30 to 1.51) | -0.76 (-2.34 to 0.81) | 0.68 |
| Carbon fibres | -0.08 (-1.39 to 1.22) | -0.63 (-1.78 to 0.52) | 0.51 |
| Ceramic fibres | 0.78 (-0.57 to 2.13) | -0.93 (-2.08 to 0.22) | 0.06 |
| Fibreglass | -.34 (-1.52 to 2.20) | -0.20 (-1.66 to 1.27) | 0.82 |
| Mineral fibres | -0.52 (-3.76 to 2.72) | 0.24 (-1.83 to 2.31) | 0.64 |

Data presented as regression coefficient (β) and 95% confidence interval (CI) unless otherwise stated. Models were adjusted for marital status, body mass index (BMI), and years of exposure to insulation works were considered as confounders. *P*^#^: p values for interaction

**Supplementary Table 6:** Effect modification by parental history of cardiac disease (no vs. yes) on the association between exposure to insulating materials and Framingham risk score.

| **Exposure** | **Framingham risk score [β (95% C)]** | | ***P^#^*** |
| --- | --- | --- | --- |
|  | **No parental history of cardiac disease (n = 455)** | **Parental history of cardiac disease (n = 169)** |  |
| Aerogels | -0.55 (-1.61 to 0.51) | 0.89 (-0.73 to 2.52) | 0.14 |
| Asbestos | 0.98 (-0.23 to 2.20) | 0.75 (-1.10 to 2.60) | 0.62 |
| Calcium silicate | -0.61 (-2.09 to 0.86) | -0.68 (-3.00 to 1.64) | 0.75 |
| Carbon fibres | -0.28 (-1.34 to 0.78) | -0.57 (-2.18 to 1.04) | 0.96 |
| Ceramic fibres | -0.02 (-1.09 to 1.06) | -0.34 (-1.99 to 1.31) | 0.92 |
| Fibreglass | 0.41 (-0.97 to 1.78) | 0.18 (-2.04 to 2.41) | 0.84 |
| Mineral fibres | 1.06 (-1.32 to 3.45) | -0.79 (-3.58 to 2.00) | 0.56 |

Data presented as regression coefficient (β) and 95% confidence interval (CI) unless otherwise stated. Models were adjusted for marital status, body mass index (BMI), and years of exposure to insulation works were considered as confounders. *P*^#^: p values for interaction
